# COVID-19 convalescent plasma donors: impact of vaccination on antibody levels, breakthrough infections and reinfection rate

**DOI:** 10.1101/2021.07.13.21260414

**Authors:** La Raja Massimo, Pacenti Monia, Grimaldi Ileana, Boldrin Caterina, Cattai Margherita, Solimbergo Erica, Battisti Anna, Scomazzon Michele, Roman Alberto, Lazzaro Anna Rosa, Vicari Stefano, Terzariol Stefano, De Silvestro Giustina

## Abstract

From April 2020 through May 2021 in Padova Province 3395 COVID-19 recovered patients were recruited as potential convalescent plasma donors and tested for SARS-CoV-2 antibodies. Since January 2021 COVID-19 vaccination campaign began in Italy, the impact of vaccination on antibody levels and suspect vaccine breakthrough infections in these subjects were investigated. Post-vaccination anti-Sars-Cov-2 antibody level in 54 previously infected subjects had an exponential increase compared to pre-vaccination level regardless of the number of vaccine doses. However after 100 days from vaccination SARS-CoV-2 antibody level tends to decline. Post-vaccination primary infections were detected in 15 cases, with 3 possible breakthrough infections after a full vaccination course. In these cases, antibody response after infection was present but weaker than the one of subjects vaccinated after natural infection. A trend toward stronger antibody response was observed with increasing distance between natural infection and vaccination. Additionally, 2 cases of asymptomatic reinfections are also discussed.

## Introduction

Since April 2020 in Padova province more than three thousand recovered patients were tested for SARS-CoV-2 antibodies to select suitable convalescent plasma-CP-donors, and more than 1000 high titre donations were collected. After anti-COVID-19 vaccination campaign took off in January 2021 some recovered donors already tested for SARS-CoV-2 antibodies after natural infection presented spontaneously for possible donations after 1 or 2 doses of COVID-19 vaccines, while other newly recruited convalescent patients reported history of COVID-19 primary infection only after one or two doses of vaccine. Additionally, follow up of CP donor cohort allowed investigation on possible cases of reinfection.

## Material and methods

All recovered COVID-19 patients that presented for CP donation in Padova province transfusion services from the first of April 2020 to the 31^st^ of May 2021 were included in the study. All participants signed informed consent at recruitment before SARS-CoV-2 antibody testing as a part of the clinical study on CP approved by the Ethics Committee of the Padova University Hospital (1). Clinical and epidemiological information was collected from patients at the time of enrolment and during following visits for plasma or blood donation. Enrolled cases were followed up for possible COVID-19 reinfection. SARS-CoV-2 infection was considered a confirmed positive RT-PCR test regardless of symptoms. Time of diagnosis was considered the date of first positive nasal and/or oropharyngeal molecular test.

Any COVID-19 vaccination reported by donors was registered during following accesses for plasma or blood donation. For the purpose of this study a vaccine breakthrough infection was defined as the detection of SARS-CoV-2 RNA in a respiratory specimen collected from a person ≥14 days after he/she has completed all recommended doses of COVID-19 vaccines (2).

Reinfection, based on epidemiological criteria, was defined as any confirmed positive RT-PCR test more than 90 days from first episode, regardless of symptoms, with at least one, negative RT-PCR tests on specimens collected between the first and second episode (3).

The LIAISON® chemiluminescence immunoassay (CLIA) SARS-CoV-2 S1/S2 IgG (DiaSorin) system was the SARS-CoV-2 quantitative antibody test utilized for donor screening. The LIAISON® CLIA values are expressed as AU (Arbitrary Units), results range between <3.8 and >400; values ≥15 AU are considered positive. According to a preliminary study ≥80 AU correlate with NA titer ≥1:160 (4). Recovered patients with CLIA results ≥80 AU were there invited to donate plasma. On CP donations an in vitro microneutralization method was used for neutralizing antibodies-NA-assessment. The cytopathic effect of the virus on VERO6 cells was evaluated in presence of diluted serum samples. After incubation the cytopathic effect by 90% has been considered to determine the titre of neutralizing antibodies (1). CP units with NA titres ≥1:160 were considered high titer units.

## Results

All COVID-19 convalescent patients that presented for CP donation from the first of April 2020 to the 31^st^ of May 2021 and were eligible for blood donation according to the Italian guidelines were included in the study. Women who had previous pregnancies and previously transfused subjects were excluded because of the risk of TRALI (Transfusion Related Acute Lung Injury) due to the possible presence of anti-HLA antibodies. In total 3395 recovered COVID-19 patients were tested for anti-SARS-Cov2 antibodies: 69.1% were males and 30.9% females, mean age was 40 (range 18-65). Median anti-SARS-CoV-2 CLIA result was 46 AU (interquartile range 23-82). Out of these 2723 were recruited before the first of March 2021 and were therefore followed up for 4 months and more from diagnosis for possible reinfections. In this group the average follow up was 240 days from diagnosis (range 455-124 days).

### Recovered and vaccinated subjects

Overall, by the 31^st^ of May 2021 54 CP donors that were tested for SARS-CoV-2 antibodies after primary infection presented for CP or blood donation after 1 or 2 doses of vaccine and were then retested for antibody level. Average age was 43 years (range 20-64) and 14 (26%) were female. According to WHO classification (5) 4 cases had severe COVID-19 disease and were admitted to hospital. 8 cases had moderate disease and 42 had mild disease, and they were managed as outpatients. On average first antibody testing from diagnosis was after 57 days (range 16-241). On average the first dose of vaccine was administered 186 days (range 41-419) after COVID-19 diagnosis. On average post-vaccination antibody test was done 50 days from first vaccination dose (range 6-169). 37 (69%) donors received one dose of vaccine: 14 Vaxzevria (AstraZeneca), 20 Comirnaty (Pfizer BioNtech) and 3 Moderna. The remaining 17 donors (31%) received two doses all of Comirnaty vaccine.

Pre and post-vaccination CLIA results were available for all 54 CP donors. In addition, both pre and post-vaccination NA titres on CP donations were available for 18 donors. Other 14 subjects donated CP only after vaccination and so only post-vaccination NA titres were available together with pre-vaccination CLIA results. Finally 8 subjects donated CP only before vaccination and so pre-vaccination NA titer was also available together with pre and post-vaccination CLIA reuslts. In the 54 CP donors pre vaccination CLIA median value was 80 AU (range <3.8-283 AU). In 25 (46%) cases CLIA values were ≥ 80 AU, the cut-off utilized to select high titre donations. In all cases after vaccination there was a robust increase in CLIA values: in 45 subjects (83%) post vaccination CLIA results were >400 AU, in 8 (15%) they were between 200 and 400 AU, and only one (2%) was between 100 and 200 AU. In 26 subjects pre-vaccination NA median titre on donation was 1:80 (range 1:20-1:320), 11 out of 26 (42%) were ≥ 1:160 and therefore considered high titre donations. Out of the 32 convalescent subjects that donated after vaccination post-vaccination NA median titre was 1:640 (range 1:1280-1:160). In details the NA titer was 1:1280 in 6 cases (19%), 1:640 in 14 cases (44%), 1:320 in 11 cases (34%) and 1:160 in one. For 18 CP donors whose pre and post-vaccination NA titres were available, the median NA titre increase was eightfold (range 1-64). There was no association between post infection antibody level or the number of doses and antibody response. However, the only two cases that had negative CLIA values after primary infection, both with <3.8 AU, the CLIA values after 2 doses of vaccine were 320 and 342 AU respectively. In eleven subjects that were vaccinated with at least one dose more than 300 days after COVID-19 diagnosis all eleven had >400 AU CLIA values and only four (36%) received 2 doses. Out of these, seven donated CP after vaccination: of these 5 (71%) had NA titre on donation of 1:1280, and two 1:640 or less. Among the 25 CP donors that were vaccinated less than 300 days after COVID-19 diagnosis only one (4%) had a NA titer of 1:1280, the remaining 24 had 1:640 or less (two-tailed Fisher exact test p<0.0001). Out of five CP donors that were tested more than 100 days after the first dose of vaccine 3 (60%) had a CLIA results < 400 AU, and all five received 2 doses of vaccine. On the contrary only 7 (14%) of the 49 subjects that were tested less than 100 days after first dose of vaccination had a CLIA result < 400 AU (two-tailed Fisher exact p=0.04), suggesting an antibody decline after this time.

### Post-vaccination infections

Overall, from the first of March 2021 to the 31^st^ May 2021, 15 candidate CP donors reported to have been infected after vaccination, 6 females and 9 males, and their mean age was 45 years (range 25-58). In 11 cases COVID-19 infection was diagnosed after the first dose of vaccine, in 4 cases after the second dose. Average time from first dose and COVID-19 diagnosis was 24 days (range 7-71 days). In all cases the infections were symptomatic and classified as mild cases. 4 subjects were vaccinated with 2 doses of Comirnaty, 5 with 1 dose of Comirnaty, 6 with 1 dose of Vaxzevria. In all subjects post infection anti-SARS-CoV-2 CLIA results were available, in 9 cases NA testing on donation was also done. Average time from diagnosis to antibody test was 43 days (range 25-66). Median CLIA value was 100 AU (range 12->400), median NA titer on donation was 1:40 (range 1:20 – >1:320).

Among 11 subjects that received only 1 dose of vaccine 7 subjects were diagnosed SARS-CoV-2 infection < 15 days from the dose and had a median CLIA value of 61 AU (range 12-203), while for the 4 CP donors that were diagnosed COVID-19 ≥ 15 days from the single dose the median CLIA value was 161 AU (range 94-331). Finally for the other 4 subjects that received 2 doses of vaccine 21 days apert before infection the median CLIA value was 289 (range 143 - >400).

Three cases, two women and one man, were classified as breakthrough infections since they occurred more than 15 days after the second dose: 31, 45 and 50 days respectively. The post infection CLIA results in these cases were 206, 372 and >400 AU respectively. NA titers on donation were 1:40, 1:160, and >1:320 respectively.

### Reinfected subjects

Out of a cohort of 2723 convalescent subjects followed up for more than 4 months only two reported microbiologically confirmed re-infection. Both, one girl and one boy, were 26 years old at the time of first diagnosis. The second infection was diagnosed respectively 201 and 347 days after the first one. Out of 1790 cumulative person-years of follow up to the estimated risk of reinfection was 1.1 × 1000 person-years. In both cases the second infection was asymptomatic and detected during contact tracing in one case and routine occupational screening in the other. One case was unvaccinated, in the second one the positive molecular sample was taken 2 days after the second dose of Comirnaty vaccine, and it was confirmed on a second sample. In the unvaccinated case after the first infection the anti-SARS-CoV-2 CLIA value was 51 AU, after reinfection it rose to 129 AU. In this case on donation after the reinfection the NA titer was 1:40. For the second case of reinfection only the NA titer was available after first infection and it was 1:40, the CLIA value after vaccination and reinfection was >400 AU.

## Discussion

For 14 months in Padova Province during COVID-19 CP donor recruitment activities more than 3000 COVID-19 patients were tested with quantitative assays for SARS-CoV-2 antibody levels. Out of the ones that were vaccinated after infection and presented again to blood collection centres for CP or blood donation we observed an exponential increase of SARS-CoV-2 neutralizing antibody levels compared to post infection levels, regardless of the type of COVID-19 vaccine, of the number of doses and of post infection antibody levels. In more than 80% of cases extremely high CLIA values - >400 AU - and NA titres - ≥1:320 – were achieved. The median increase of NA was eightfold higher than after natural infection. These data confirm recent published reports that also a single dose of vaccine is able to elicit an robust increase of SARS-CoV-2 antibody titre (6,7) in previously COVID-19 infected patients. Our data additionally suggest that a longer interval between infection and vaccination, in our case more than 300 days, is associated to a stronger booster effect on antibody levels.

Infection after vaccination was not uncommon in convalescent subjects under study, it occurred mainly during the first four weeks after the first dose and it decreased after the second as already reported (7). In our observation antibody response after post-vaccination infection appears dependent on the time-lag between infection and first vaccination dose, increasing over time. Among the observed cases of post-vaccination infections, the SARS-CoV-2 antibody levels were on average higher than natural infection but lower than subjects vaccinated after natural infection. We identified three breakthrough infections among out CP donors, i.e. after 2 weeks from the second dose of vaccine. All of them were symptomatic mild infections and again the SARS-CoV-2 antibody level appears to rise when time-lag between two events increases. It was observed that novel SARS-CoV-2 variants may be associated with breakthrough infections (8,9), we couldn’t confirm this in our study since genetic typing on viral isolates was not available.

Reinfection in the population under observation was exceedingly rare: only two cases and both asymptomatic. In both cases SARS-CoV-2 antibodies were present after first infection although at a low level. In our experience reinfection rate appears lower than previously reported (11). However, since reinfections were incidentally detected during occupational screening and contact tracing activities, and not as a result of systematic active microbiological follow up program, it is likely that during outbreaks short lived unnoticed reinfections are more frequent than what was observed in our study. Our limited data, only one case, show that in unvaccinated subjects asymptomatic reinfections are associated with increase in antibody level as determined by CLIA.

In conclusion this study of convalescent plasma donor cohort offered an opportunity to observe SARS-CoV-2 antibody response of COVID-19 infected subjects between 18 and 65 years of age and the impact of vaccination campaign on antibody levels in these subjects.

## Data Availability

All data are available in the Padova transfusion department database and Padova University Hospital Microbiology and Virology service information system

## Notes

### Competing Interest Statement

The authors have declared no competing interest.

### Clinical Trial

This is an observational study on convalescent plasma donors as part of the Study Protocol on Covalescent Plasma that has been approved by the Ethics Committee (EC) of the Padova University Hospital, Details on the protocol were already described elsewere https://www.ncbi.nlm.nih.gov/pmc/articles/PMC8103741/

### Funding Statement

Funding of convalescent plasma activities was provided by the Regione del Veneto.

### Author Declarations

The present study is a part of the study protocol : Interventional study to evaluate the efficacy of treatment with plasma, collected from recovered COVID-19 patients (convalescent plasma or hyperimmune plasma), in patients with severe COVID-19 infection, authorised by the ethics committee of the hospital of Padua, the Italian National Blood Centre, and the Veneto region

